# Street Survival to Substance Use: The Impact of Drug Addiction on Street Children in Dhaka, Bangladesh

**DOI:** 10.1101/2025.10.07.25337500

**Authors:** Nigar Sultana Sharmi, Maliha Mahazabin

**Affiliations:** Department of Child Development and Social Relationship, University of Dhaka, College of Home Economics (Constituent), Azimpur, Dhaka 1205, Bangladesh; Canadian Trillinium School, Gulshan – 2, Dhaka; Statistics Discipline, Science Engineering and Technology School, Khulna University, Khulna – 9208, Bangladesh

**Keywords:** Street Children, Drug Addiction, Substance Abuse, Child Vulnerability, Dhaka City, Malnourished

## Abstract

Bangladesh has one of the largest populations of street children in South Asia, with millions growing up without stable housing, food security, or parental care. A significant proportion is concentrated in Dhaka, where exposure to poverty, neglect, and negative peer influence creates an environment in which drug use becomes a common coping strategy. Substance use among these children not only reflects immediate survival mechanisms but also leads to profound physical, psychological, and social vulnerabilities, underscoring the urgent need for research and intervention. The study conducted in Dhaka city and collected data were organized and analyzed using descriptive statistical methods. Our study found that about 40% of the surveyed street children are currently using drugs. Alarmingly, 33.3% of them could not recall their family members or had no memory of their familial background. Children between the ages of 6 and 9 appeared to be the most vulnerable, with nearly 70% in this age group becoming addicted to drugs. As a result, approximately 40% of the children reported experiencing physical, mental, emotional, or social problems linked to substance use. Additionally, 30% of the children who had previously attended school had since dropped out and were no longer receiving formal education. Our findings also reveal that 43.33% of the children initiated drug use through cigarettes. When asked about potential solutions, 73.33% of the respondents agreed that a child’s own willingness plays the most critical role in overcoming addiction. These findings highlight the urgent need for targeted interventions addressing the root causes of drug addiction among street children in Dhaka. Comprehensive rehabilitation programs, early childhood support, nutritional assistance, and community-based outreach are critical to reducing substance abuse in this vulnerable population. Policymakers, NGOs, and health authorities must prioritize long-term, child-centered strategies to ensure protection, recovery, and social reintegration for street children in Bangladesh.

## 1. Introduction

“Today’s children, tomorrow’s future” is a phrase often quoted in textbooks and educational discourse. However, in the context of Bangladeshi society, the actual outcomes are far from ideal when we look into the real situation of many children in this country. Particularly among street children and disadvantaged youth, we see a very different, often worse, reality. They grow up neglected and unwanted by society as well as lack not only education, but also sufficient food and other basic necessities **[1]**. An alarming picture of neglect, vulnerability, and marginalization, many fall into bad company and become involved in anti-social activities. Among these, substance use or drug taking is one of the most significant negative outcomes, imposing serious consequences on the wider social system **[1–3]**.

Street children in Bangladesh grow up in an environment where they are widely regarded as unwanted and disregarded by both the public and the state **[1,4]**. They are deprived not only of formal education but also of access to sufficient food, shelter, and healthcare. Without even one basic need fully met, these children experience chronic insecurity, hopelessness, and exclusion. As a result, a growing number of children are turning to drug or substance use. Drugs and psychoactive substances are not used for medical purposes in this context; rather, they serve as addictive agents that alter consciousness and behavior. The use of drugs among children can severely impair cognitive functions, damage emotional regulation, and lead to sudden, unpredictable behavioral changes. Substances commonly used by Bangladeshi street children include alcohol, heroin, opium, marijuana, marijuana derivatives (such as majurina), yaba, and, perhaps most alarmingly, even cigarettes and inhalants like glue (“dandy”) **[5,6]**. According to the Bangladesh Children Rights Forum, around 85% of street children are suffering from some form of substance use. A systematic review of 50 studies from 22 countries found that lifetime substance use among street children is high (pooled prevalence ∼60%), with inhalant use being one of the predominant forms (∼47%) in many regions **[7]**.

According to the Australian Government Department of Health and Aged Care, drugs can be classified based on their effects on the body **[8]**: Depressants: Slow down the function of the central nervous system; Hallucinogens: Affect sensory perception and alter the way individuals see, hear, taste, smell, or feel; Stimulants: Speed up the function of the central nervous system. These substances, particularly inhalants and cigarettes, are often cheap and readily available, making them easily accessible to children living on the streets. This accessibility, combined with a lack of supervision and awareness, leads to widespread use and addiction **[4]**.

Drug addiction among street children not only destroys their individual potential but also poses a serious threat to societal well-being. Many of these children lack any understanding of the long-term consequences of substance use or the laws designed to prevent it **[7]**. They often perceive only the punitive side of legal and societal responses, without grasping the protective or rehabilitative aspects. This disconnect may contribute to increased aggression, depression, defiance, and social withdrawal, further deepening their marginalization. Street children represent one of the most vulnerable and high-risk groups in the country when it comes to drug abuse **[3,5,9]**. The combination of poverty, lack of education, and unmet basic needs pushes them toward curiosity, risky behavior, and ultimately addiction. Once entrapped in this cycle, many are lost to the “darkness” of substance dependency, often beyond recovery, unless urgent and effective intervention is provided **[9]**.

In recent years, the use of substances and illicit drugs has become increasingly widespread, with rates continuing to rise day by day. Our research investigates how street children in Dhaka become involved in drug addiction, where and how they access these substances, and who may be facilitating their use. Drug addiction among this vulnerable group not only affects their individual lives but also has broader implications for the social fabric and urban stability of Dhaka, the capital of Bangladesh.

### Objectives of the Study

This study aims to explore the following key questions:

- Why do street children become addicted to drugs?
- To what extent are they aware of the negative consequences of drug use?
- What are the main sources through which they obtain drugs?
- What are the physical health impacts of drug use on these children?
- What are the psychological or mental health consequences they experience?
- How do they perceive society’s response or feedback toward them as drug users?
- Does the widespread drug use among street children contribute to broader societal vulnerability and harm?
- What is the general perception or attitude of society toward street children involved in drug use?

## 2. Materials and methods

### Study Design and Setting

This research employed a cross-sectional, descriptive design to investigate the consequences of drug addiction among street children in Dhaka, the capital city of Bangladesh. The study was conducted across several major urban areas where street children are commonly found. These locations included Mirpur, Azimpur, Mohammadpur, Uttara, Suhrawardy Udyan, Kamalapur, and Shaheed Minar—all of which are known for their high population of street children and informal settlements.

### Participants and Sampling

A total of 30 street children, aged between 7 and 15 years, were selected to participate in the study. Participants were approached directly in the street, parks, or nearby temporary shelters. A purposive sampling method was used to ensure that only children with a history of drug use or addiction were included, as identified through initial screening and informal interviews.

### Data Source

Data were collected using a structured survey questionnaire, which included both open-ended and close-ended questions to allow for both quantitative analysis and deeper insights. The questionnaire was developed in Bangla for ease of understanding and covered multiple dimensions including demographic information, sources of obtaining drugs, physical and mental health symptoms. The questions were administered orally by the researchers, given the low literacy levels of participants. Responses were recorded manually and transcribed for analysis. All data used in this study were primary, collected directly from the participants during field visits between 5^th^ September and 27^th^ November 2019.

### Data Analysis

Collected data were organized and analyzed using descriptive statistical methods. Frequencies and percentages were calculated to describe the distribution of drug use, health effects, and other categorical variables. Statistical software was utilized for analysis purpose.

### Ethical Considerations

This study was conducted in accordance with the ethical standards outlined in the Declaration of Helsinki. Ethical approval was obtained from the Institutional Review Board (IRB) prior to data collection. Due to the participants being street children, most of them were without legal guardians; a waiver of parental consent was granted by the IRB, recognizing the unique vulnerability and inaccessibility of this population. Informed verbal consent was obtained from each child after explaining the study objectives, procedures, and confidentiality assurances in age-appropriate language. Additionally, necessary permission was obtained from relevant local authorities to conduct research in the selected areas. Children were assured that their responses would remain anonymous and confidential. No identifying information was collected. The research did not include any form of intervention or treatment. All participation was voluntary, and participants could withdraw at any time.

## 3. Results

The findings indicate that family connection and awareness remain limited among street children who use drugs (Table 1). Only half (50%) maintained some form of contact with their families, while a third (33.3%) could not recall their family at all, reflecting severe disconnection from parental or kinship ties.

**Table 1.** Background, Drug Use Patterns & Connections.

| Variables | Frequency (n) | Percentage (%) |
| --- | --- | --- |
| **Connection / Contact with Family** |  |  |
| Unable to recall any connection with family | 10 | 33.3% |
| Have connection / contact | 15 | 50.0% |
| Former contact | 5 | 16.66% |
| **Who in family knows about your drug use** |  |  |
| Parents | 7 | 23.3% |
| Siblings | 2 | 6.67% |
| Nobody | 19 | 63.3% |
| Others | 2 | 6.67% |
| **Do other family members use drugs** |  |  |
| Currently take drugs | 12 | 40.0% |
| Took once | 2 | 6.67% |
| Not take | 9 | 30.0% |
| Don't know | 7 | 23.33% |
| **Family's reaction / perception when you take drugs** |  |  |
| Scolding | 4 | 13.33% |
| Punishment | 4 | 13.33% |
| Nothing | 8 | 26.67% |
| Forbid to take | 14 | 46.67% |
| **Age when respondent first took drugs** |  |  |
| 6–9 years | 21 | 70.0% |
| 10–13 years | 8 | 26.67% |
| 14–17 years | 1 | 3.33% |
| **Duration of drug use (years)** |  |  |
| 1 to 3 | 15 | 50.0% |
| 3 to 5 | 8 | 26.67% |
| 5 to 10 | 8 | 26.33% |
| **Schooling status** |  |  |
| Yes, went to school | 10 | 33.33% |
| Never attended | 10 | 33.33% |
| Once but dropped out | 9 | 30.0% |
| Not willing / no interest | 1 | 3.33% |
| **Work / Hard Labor Hours** |  |  |
| Work all day | 17 | 57.0% |
| If someone gives work | 12 | 40.0% |
| No work / no hard labor | 1 | 3.0% |
| **Type of drug(s) taken so far** |  |  |
| Cigarette | 13 | 43.33% |
| Dandy (glue/inhalants) | 12 | 40.0% |
| Other substances | 5 | 16.67% |
| **Most-used drug type** |  |  |
| Cigarette | 19 | 63.33% |
| Dandy | 5 | 16.67% |
| Other | 6 | 20.0% |
| **Drug usage frequency per day** |  |  |
| As many times as possible | 21 | 70.0% |
| One time | 6 | 20.0% |
| Not daily / occasional | 3 | 10.0% |
| **Interval between uses** |  |  |
| Daily | 22 | 72.0% |
| After 1–2 days | 5 | 15.0% |
| After 2–3 days | 3 | 11.0% |
| Any time / no fixed pattern | 1 | 2.0% |
| **Time of drug use** |  |  |
| Day time | 3 | 10.0% |
| Night time | 4 | 13.33% |
| No fixed time / anytime | 23 | 76.67% |
| **Amount spent per use** |  |  |
| 10–30 taka | 14 | 46.67% |
| 50 taka | 11 | 36.67% |
| 100 taka | 4 | 13.33% |
| 200 taka | 1 | 3.33% |
| **What they perceive is most critical to quitting** |  |  |
| Own willingness | 22 | 73.33% |
| Family support | 4 | 13.33% |
| Rehabilitation / treatment | 4 | 13.33% |

Disclosure of drug use within the family was minimal: nearly two-thirds (63.3%) reported that no family member knew about their drug involvement, with parents’ awareness reported by 23.3% and siblings’ by only 6.7%. A considerable proportion (40%) indicated that other family members were also drug users, suggesting potential intergenerational or household-level patterns of substance abuse. Family responses to drug use varied: 46.7% said their families forbade use, while others reported either indifference (26.7%) or negative disciplinary responses such as scolding (13.3%) or punishment (13.3%).

Initiation into drug use occurred alarmingly early, with 70% beginning between ages 6 and 9. In terms of duration, half reported using for 1–3 years, while roughly equal proportions had used for 3–5 years (26.7%) or 5–10 years (26.7%). Educational attainment was poor: only 33.3% had ever attended school, with 30% dropping out, and 33.3% never enrolling. Labor participation was also precarious; 57% worked full days, while 40% only worked when offered temporary opportunities. Cigarettes (43.3%) and “dandy” (40%) were the most common substances, with cigarettes being the most frequently consumed (63.3%). Use was intensive, with 70% reporting multiple uses per day, 72% using daily, and 76.7% lacking fixed use times.

Financially, nearly half (46.7%) spent 10–30 Bangladeshi taka per use, while 36.7% spent about 50 taka. Regarding quitting perceptions, the majority (73.3%) believed that personal willingness was the most crucial factor, whereas family support and rehabilitation were each mentioned by 13.3%.

A significant proportion (43.3%) acknowledged that their dosage was increasing, while 40% reported no change (Figure 1).

**Figure 1.**
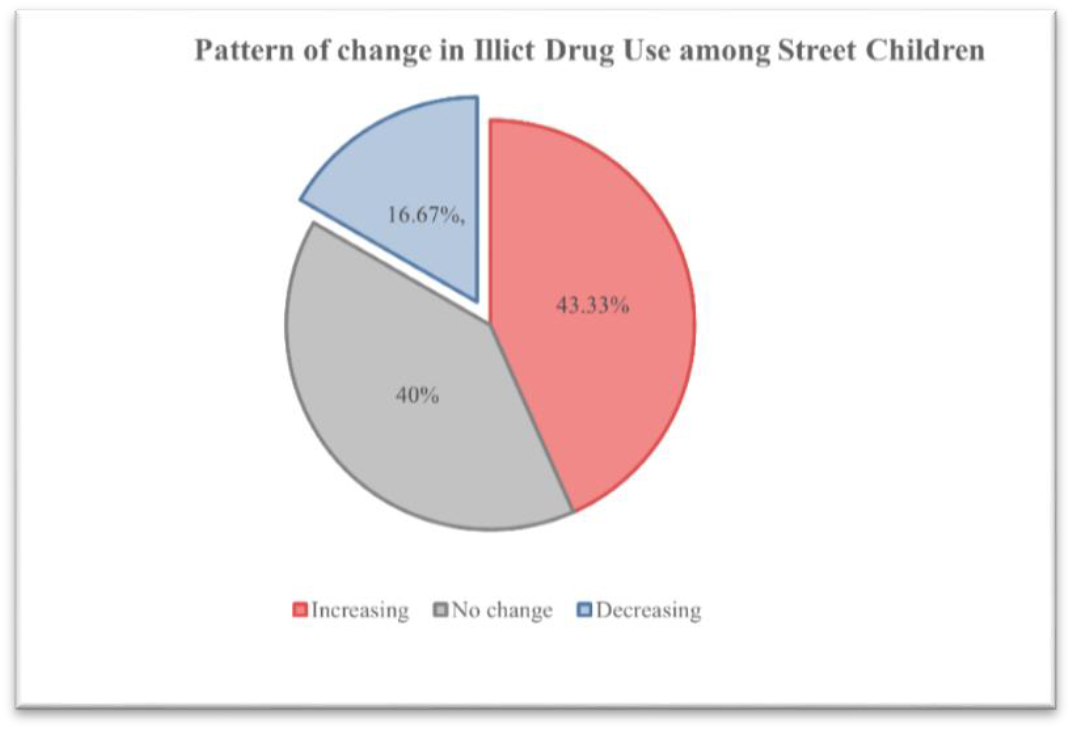
Reported Changes in the Frequency of Illicit Drug Use by Street Children

The study revealed several key factors contributing to drug addiction among street children in Dhaka. A notable proportion of participants (16.7%, n = 5) reported coming from households where one or both parents were addicted to alcohol or drugs (Table 2).

**Table 2.** Reasons behind drug addiction among street children.

| Reason | Frequency (n) | Percentage (%) |
| --- | --- | --- |
| Addicted/alcoholic parents | 5 | 16.7% |
| Easy availability of substances | 10 | 33.2% |
| Peer pressure / bad company | 9 | 30.0% |
| Emotional coping (hunger/stress) | 6 | 20.1% |

In households where parents were alcoholic or addicted, children often witnessed substance use from an early age. This exposure normalized drug-taking behavior, making them more likely to experiment themselves. Another major factor reported was the easy availability and low cost of drugs such as cigarettes, glue (inhalants). Many children indicated that these substances could be obtained from local vendors, peers, or even other street children.

Peer influence also played a significant role in initiating drug use. About 30% of participants stated that they began using drugs due to pressure or encouragement from friends, especially older children in the street environment. In addition to peer pressure and family influences, many children reported turning to drugs as a way to cope with the emotional and physical hardships of life on the street. These included extreme hunger, fear of violence, loneliness, sadness, and hopelessness. Some children mentioned that taking substances like glue or cigarettes helped them “forget hunger”.

Table 3 presents findings on how street children in Dhaka view and understand the negative consequences of drug use. A majority (60%) believe that taking drugs is “bad,” showing a broadly negative attitude. A small fraction (6.67%) think drug use is “good,” and about one-third (33.33%) are unsure or neutral. This indicates that although many know it is harmful, a fair number are ambivalent or lack strong conviction.

**Table 3.** Awareness of negative consequences of drug use.

| Measure | Frequency (n) | Percentage (%) |
| --- | --- | --- |
| **Opinion of taking drugs** |  |  |
| Think it is “good” | 2 | 6.67% |
| Think it is “bad” | 18 | 60% |
| Other / unsure | 10 | 33.33% |
| **Knowledge of negative consequences** |  |  |
| Know bad side of taking drugs | 26 | 86.67% |
| Do not know | 2 | 6.67% |
| Do not want to know | 2 | 6.67% |
| **Intentions whether to continue or stop** |  |  |
| Will continue using | 8 | 26.67% |
| Will stop using outright | 7 | 23.33% |
| Will stop if they get support from others | 11 | 36.67% |
| Other / unsure | 4 | 13.33% |
| **Attitude toward whether others should take drugs** |  |  |
| Want others to take drugs | 7 | 23.33% |
| Do not want others to take drugs | 17 | 56.67% |
| Do not know / unsure | 6 | 20% |

Most participants (86.67%) report that they know some of the bad consequences of drug use. A small minority (6.67%) say they do not know, and another 6.67% say they “do not want to know,” which suggests some avoidance or denial. These responses suggest that knowledge is fairly high, though depth and correctness of that knowledge likely vary. Intentions About Future Use, the responses are mixed: 26.67% intend to continue use, 23.33% say they will stop outright, and 36.67% indicate they would stop if they received support from others. A minority (13.33%) are unsure or have “other” responses. This suggests that while knowledge exists, many feel they cannot quit alone and need external support (social, medical, familial). Attitude toward others using drugs, 56.67% do not want others to take drugs, reflecting concern or empathy. However, 23.33% say they want others to use drugs, and 20% are unsure. The fact that almost one-quarter encourage or accept others’ drug use suggests normalization, peer pressure, or perhaps identification with drug users, which may reinforce usage.

Table 4 demonstrates findings from street children (aged 7 15) in Dhaka regarding how they obtain drugs, how they first began drug use, whether they sell or deliver drugs, ways they get money to procure drugs, and who helps them in procurement.

**Table 4.** Sources of Drug Access & Initiation among Street Children.

| Variables | Frequency (n) | Percentage (%) |
| --- | --- | --- |
| **Who they get drugs from** |  |  |
| Family | 1 | 3.33% |
| Friends | 9 | 30% |
| Collect by themselves | 20 | 66.67% |
| **How they first took drugs** |  |  |
| Self-curiosity | 17 | 53.67% |
| Provocation by others | 10 | 33.33% |
| Seeing friends use | 3 | 10% |
| **Selling drugs** |  |  |
| selling by self | 4 | 14% |
| Not selling | 22 | 75% |
| Would sell if opportunity arises | 3 | 10% |
| Others | 1 | 1% |
| **Selling to/delivering to others** |  |  |
| Delivering to friends | 14 | 46.67% |
| Not delivering or selling | 14 | 46.67% |
| Other / unsure | 2 | 6.67% |
| **Ways of getting money to buy drugs** |  |  |
| Theft or hijacking | 3 | 10% |
| Begging | 5 | 16.67% |
| Taking loans | 2 | 6.67% |
| Other sources | 20 | 66.67% |
| **Who helps them procure drugs** |  |  |
| Family members | 8 | 26.67% |
| Friends | 20 | 66.67% |
| Others | 2 | 6.67% |

Self-collection is the predominant way children access drugs as about two-thirds say they get the substances themselves, rather than relying on family or friends. Friends also play a major role both as a source (30%) and as helpers in procurement (66.67%). Moreover, family involvement is relatively low (3.33%) in directly giving drugs, but about one in four children report that family members have helped them procure drugs. Initiation to drug use is most often driven by self-curiosity (53.67%), followed by peer provocation (33.33%). Seeing friends use contributes less (10%). A small share is involved in selling drugs themselves (14%), and nearly half have delivered or given drugs to friends (46.7%). To buy drugs, many street children rely on “other” sources (66.7%), such as collecting or selling recyclable materials, doing day labor, or petty work, followed by begging (16.7%), theft or hijacking (10%), and taking loans (6.7%) to raise money.

A majority of street children reported feeling good during their first experience with drugs (54%), while 44% felt bad (Table 5). This initial positive feeling may contribute to the continuation of drug use.

**Table 5.**
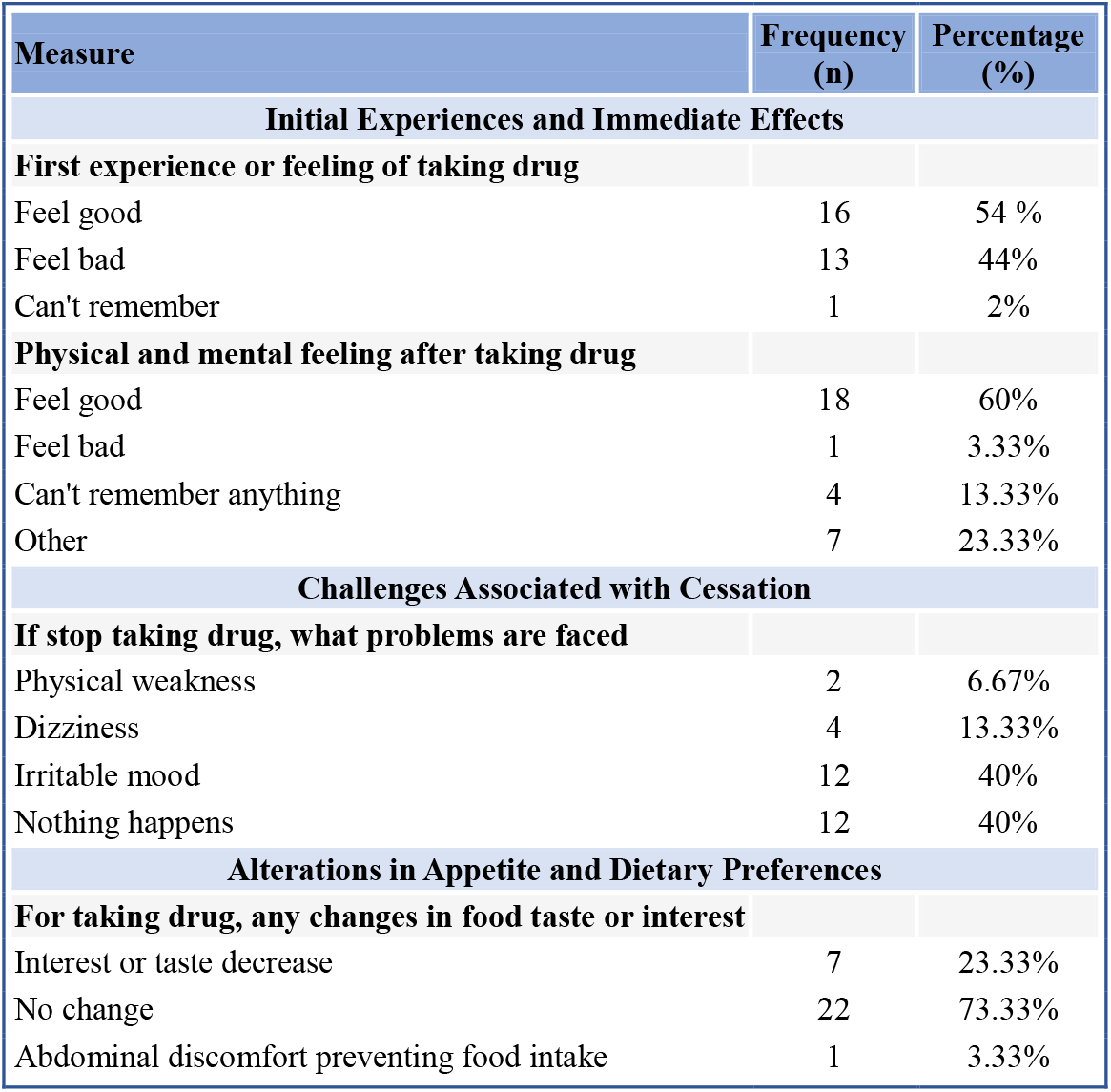
Physical and Psychological Health Impacts of Drug Use among Street Children.

After taking drugs, 60% reported feeling good, 3.33% felt bad, and 13.33% couldn’t remember their experience. The remaining 23.33% reported other unspecified effects. When attempting to stop drug use, 40% experienced irritability, 13.33% dizziness, and 6.67% physical weakness. However, 40% reported no noticeable changes, indicating variability in withdrawal symptoms. A significant portion (73.33%) reported no change in their interest or taste in food, while 23.33% experienced a decrease in appetite. Only 3.33% reported abdominal discomfort that prevented them from eating food.

### Perception of Society’s Response Toward Street Children Who Use Drugs

The majority of participants in this study reported feeling rejected, ignored, or verbally abused by members of the public due to their drug use. Many children expressed that they are often labeled as “addicts,” “criminals,” or “bad children,” and treated with disdain and avoidance in public places. Majority of the surveyed children reported that people avoid or scold them when they are seen using or asking for drugs, while few said that passersby sometimes offer them food or minor support but do not engage further.

### Perceived Impact of Drug Use on Society and Social Vulnerability

When asked whether they believe their drug use affects society, most children were either unaware or did not fully understand the societal consequences. However, through observation and further questioning, it became clear that community safety, public health, and urban security are negatively affected by the widespread visibility and behaviors of drug-using street children. Most of children acknowledged that people fear them or avoid certain areas where many drug-using children gather (e.g., train stations, parks, markets).

### Society’s General Attitude Toward Street Children and Drug Users

When asked how they think society views them overall, most children responded with phrases like “bad people,” “garbage,” “nobody wants us,” or “people hate us.” Majority reported feeling socially excluded and believed that people consider them dangerous or unworthy of help. Only few respondents believed that the public sees them as “children in need,” and even fewer felt that they were treated as equal to other children.

## 4. Discussion

The impact of drug addiction among street children in the 21st century is profound, with far-reaching consequences for both individual well-being and broader society. While international observances such as the International Day Against Drug Abuse on 26th June raise awareness, celebrating the day alone is insufficient to address the pervasive challenges faced by street children **[10]**. The findings of this study highlight the multidimensional vulnerabilities of street children in Dhaka, particularly regarding substance use. The present study examined drug addiction among street children in Dhaka, Bangladesh, exploring the factors contributing to substance use, awareness of its consequences, sources of drugs, health impacts, and societal perceptions. The findings reveal a complex interplay of individual, familial, and structural vulnerabilities, consistent with regional and global research on marginalized juvenile populations.

### Familial Detachment and Early Vulnerability

Approximately one-third of respondents reported limited or no contact with their families, reflecting profound disconnection from parental structures. This finding aligns with prior studies documenting abandonment and weak kinship ties among urban street children in Bangladesh. Such lack of familial attachment has been identified as a key driver of substance use in marginalized populations **[7,9,11]**. Moreover, significant amount of children reported other family members also using drugs, indicating potential normalization of substance use within certain household contexts **[12]**.

### Educational Deprivation and Socioeconomic Challenges

Educational deprivation was evident among the participants, with only a third having ever attended school. This lack of educational opportunities significantly contributes to the cycle of vulnerability among street children. Additionally, irregular and precarious labor participation reflects broader structural exclusion from stable livelihoods, reinforcing substance use as both a coping mechanism and a survival strategy **[13]**.

### Drivers of Drug Addiction Among Street Children

The study found that street children become addicted to drugs due to multiple factors, including familial detachment, peer influence, and coping with adverse circumstances such as hunger, fear, and insecurity. About one-third of participants reported minimal or no contact with family, highlighting profound parental disconnection. This lack of familial attachment has been previously identified as a key risk factor for substance use among marginalized children **[12,14]**. Peer modeling and the normalization of drug use within street networks were also significant contributors, aligning with prior studies showing that social belonging and imitation strongly influence early initiation of substance use **[15]**.

### Awareness of Negative Consequences

Despite high exposure to drugs, many children demonstrated limited understanding of the associated health risks. While some expressed a desire to quit upon learning about adverse effects, misinformation, often transmitted through peers or older street children, remained prevalent. This aligns with previous research indicating that street-involved youth often lack access to accurate health education and harm reduction programs **[16]**.

### Sources and Accessibility of Drugs

Self-procurement from street dealers was the primary method of obtaining substances, consistent with reports from the Department of Narcotics Control, Bangladesh **[17]**. Drugs such as cigarettes and inhalants (“dandy”) were readily accessible due to their low cost, reinforcing early initiation and habitual use **[3]**.

### Physical Health Impacts

Substance use significantly affected children’s physical health. Respondents reported alterations in appetite, poor nutrition, and physical weakness, especially during withdrawal episodes. Early initiation of inhalants and other substances is associated with neurocognitive deficits, growth retardation, and long-term physiological consequences **[3,8,11]**.

### Psychological and Mental Health Consequences

Drug use served as a coping mechanism for emotional trauma, including anxiety, fear, and social isolation. However, continued consumption led to irritability, aggression, memory deficits, and dependency, reflecting the psychological burden of addiction. These findings are consistent with studies demonstrating high rates of mental health issues among street-involved drug users **[3,7,17]**.

### Patterns of Drug Use and Dependency

The most frequently used substances among the children were cigarettes and inhalants such as “dandy,” reflecting patterns reported in Bangladesh and other South Asian settings where affordability and easy accessibility drive consumption **[3,9]**. The study found that many participants used substances multiple times a day, with some gradually increasing their dosage, emphasizing the chronicity of use and the development of dependency-level behaviors **[6,10]**.

### Perceptions of Treatment and Rehabilitation

Importantly, the majority of participants believed that their own willingness was the key factor for quitting, while only a small fraction highlighted the role of family support or rehabilitation. This perception indicates both a lack of awareness of structured treatment services and mistrust in institutional interventions. Previous studies have noted similar trends, suggesting that combining individual motivation with accessible rehabilitation and reintegration programs yields more sustainable recovery outcomes **[2,12]**.

### Social Stigma and Marginalization

The study also highlighted the social stigma faced by street children, with many reporting negative perceptions from society. experiences of stigmatization, physical abuse by authorities, and negative attitudes from the public. Many children believed society views them as a burden, reinforcing feelings of hopelessness and social exclusion. Prior literature similarly documents the role of stigmatization in perpetuating vulnerability and impeding rehabilitation **[11,12,17]**. This stigmatization can further marginalize these children, pushing them deeper into drug dependency. Previous research has shown that public attitudes toward street children often associate them with delinquency and crime, exacerbating their vulnerability **[3,4,14]**.

### Broader Societal Vulnerability

Widespread drug use among street children contributes to broader societal challenges, including public safety concerns, urban disorder, and strain on social services. The combination of early initiation, chronic use, and marginalization perpetuates cycles of poverty, delinquency, and public health risks, emphasizing the societal dimension of this problem **[2,4,11]**

### Limitations and Future Research

This study provides important insights into drug addiction among street children in Dhaka; however, several limitations should be noted. The sample was relatively small and drawn from specific urban areas, which may limit generalizability to all street children in Bangladesh. Data were primarily self-reported, which may introduce biases, and the cross-sectional design prevents causal inferences regarding family factors, drug use initiation, and health outcomes. Additionally, formal psychological assessments were not conducted, limiting precise measurement of mental health conditions, and contextual factors such as local interventions or community attitudes may have influenced responses.

Future research should employ larger, more representative samples and consider longitudinal designs to examine the long-term effects of early drug use. Incorporating standardized mental and physical health assessments would provide more accurate evaluation of outcomes, and studies assessing the effectiveness of family-based interventions, harm-reduction programs, and community support initiatives could inform evidence-based policies. Such research would help develop targeted strategies to mitigate addiction and improve the well-being of street-involved children.

## 5. Conclusion

This study highlights the multifaceted reasons behind drug use among street children in Dhaka, including individual, familial, and social factors, and underscores its physical and psychological consequences. Despite some awareness of the negative effects of drugs, knowledge alone does not consistently translate into behavior change, as attitudes and intentions vary among children. Peer influence and normalization of substance use within social networks further reinforce risky behaviors, and a subset of children even encourages others to use drugs, reflecting deep vulnerability and despair. Findings emphasize the need for comprehensive interventions that combine awareness campaigns, rehabilitation, counseling, and peer-support programs. Governmental and non-governmental organizations must ensure that street children’s basic needs, rights, and access to services are met, while broader societal and community-level efforts, including drug regulation, social awareness, and promotion of ethical and religious education, are essential to reduce substance use and its associated harms.

### Implications and Recommendations

Effective interventions require a multi-pronged approach:

- Family reintegration and support to reduce vulnerability.
- Educational and harm-reduction programs to address misinformation, spread awareness, and provide coping strategies.
- Accessible rehabilitation services tailored for street children, combining medical, psychological, and social support.
- Community engagement to reduce stigma and facilitate social reintegration.

Such strategies, implemented collaboratively by government, NGOs, and community stakeholders, can mitigate the long-term consequences of drug addiction and promote healthier developmental trajectories.

## Data Availability

All data produced in the present work are contained in the manuscript.

## Author Declarations

This work did not receive any funding. The authors declared no competing interests.

All relevant ethical guidelines have been followed; the study received approval Ethical Clearance Committee review board to ensure compliance with ethical principles and safeguard the rights of participants.

